# Linking PI-2620 tau-PET, fluid biomarkers, MRI, and cognition to advance diagnostics for progressive supranuclear palsy

**DOI:** 10.1101/2024.10.14.24315486

**Authors:** Roxane Dilcher, Charles B. Malpas, William T. O’Brien, Stuart J. McDonald, Craig Despott, Kelly Bertram, Matthew P. Pase, Meng Law, Terence J. O’Brien, Lucy Vivash

## Abstract

**Importance:** Identifying biomarkers for primary tauopathies such as progressive supranuclear palsy (PSP) is crucial for improving diagnosis and establishing clinical trial and treatment endpoints.

**Objective:** To determine whether brain tau deposition, measured by ^18^F-PI-2620 tau-PET, is associated with fluid biomarkers, regional atrophy, and cognition in PSP. We hypothesized that increased subcortical tau uptake correlates with elevated fluid biomarkers and explored its relationship with brain volume and cognition, providing insights into disease progression.

**Design:** Cross-sectional analysis of baseline data from the SEL003 clinical trial of sodium selenate for PSP, conducted from July 2021 to August 2024.

**Setting:** Multicentre study across six clinical sites in Australia.

**Participants:** 28 patients with probable PSP (Richardson’s syndrome) and 11 age- and sex-matched healthy controls from the Monash University Brain and Cognitive Health (BACH) study. Patients were over 40 years old, had symptom onset within five years, and had no structural abnormalities on MRI.

**Exposures:** ^18^F-PI-2620 tau-PET, MRI, and cognitive testing were conducted in all patients. A subset of 24 patients and 10 healthy controls had blood and CSF samples analysed for NfL, GFAP, and t-tau.

**Main Outcome and Measures:** Associations between tau-PET uptake, fluid biomarkers (NfL, GFAP, t-tau, NfL/t-tau, GFAP/t-tau, GFAP/NfL), brain volume, and cognition. Secondary outcomes included group differences (patients vs controls) in fluid biomarkers. Standardized uptake value ratio (SUVr) values were calculated using cerebellar grey matter as the reference. Cognitive testing was performed using a battery of tests.

**Results:** Among 28 patients (median [IQR] age, 66 [62-70] years; 43% females), subcortical tau uptake was significantly associated with elevated NfL/t-tau (β=0.01; 95% CI, -0.01-0.02), reduced GFAP/NfL (β=-0.02; 95% CI, -0.01--0.01), reduced brain volume, and executive function impairment. Patients exhibited higher fluid biomarker levels in both plasma and CSF compared to controls (median [IQR] age, 65 [62-68] years; 60% female), particularly in biomarker ratios (NfL/t-tau: 98.9%, 95% CI 96.16%-100%).

**Conclusions and Relevance:** These findings highlight the utility of ^18^F-PI-2620 as an in-vivo biomarker of tau pathology in PSP, with fluid biomarkers serving as valuable surrogate markers. Integrating both could improve diagnostic accuracy and treatment evaluation.

**Trial Registration:** Australian New Zealand Clinical Trials Registry (ACTRN12620001254987).

**Key points:** **Question:** Is tau accumulation, as measured by ^18^F-PI-2620 tau-PET, associated with fluid biomarkers, brain volume, and cognitive function in patients with progressive supranuclear palsy (PSP)?

**Findings**: In this cross-sectional study of 28 patients with probable PSP, subcortical tau uptake correlated positively with NfL levels and NfL/t-tau ratios, and negatively with GFAP levels and GFAP/NfL ratios in CSF and plasma. Tau-PET was associated with lower brain volume and cognitive impairments.

**Meaning:** These findings suggest that ^18^F-PI-2620 tau-PET is a valuable tool for assessing tau pathology and its relationship with neurodegeneration and clinical outcomes in PSP. Fluid biomarker ratios may serve as surrogate markers in future clinical trials.

## Introduction

Tauopathies are neurodegenerative diseases characterized by the pathological accumulation of tau protein aggregates in the brain, leading to distinct patterns of neurodegeneration and symptom progression^1–4^. Progressive supranuclear palsy (PSP) is a primary tauopathy, marked by midbrain atrophy and tau deposition in neurons and glial cells, predominantly in the brainstem and basal ganglia. Clinically, PSP presents as an atypical parkinsonian syndrome with gait instability, frequent falls, oculomotor, and executive dysfunction^5^.

Currently, the probable diagnosis of PSP relies on clinical criteria^6^, with the definitive diagnosis confirmed only post-mortem. The lack of disease-specific imaging or fluid biomarkers hinders insights into the pathophysiology and progression of the disease, and therefore early diagnosis and targeted treatment development.

The second-generation tau-PET tracer ^18^F-PI-2620 has shown promise due to its selective affinity for 4R-tau and 3/4R-tau deposits, demonstrated through autoradiography and immunohistochemistry studies and improved off-target binding profile compared to earlier-generation tracers^7^. It has the potential to detect tau pathology across various tauopathies, including PSP, corticobasal syndrome (CBS), and Alzheimer’s disease (AD). In vivo studies have revealed elevated ^18^F-PI-2620 binding in the basal ganglia of PSP patients, differentiating it from the cortical tau distribution typically seen in AD^8,9^. Existing fluid biomarkers, such as neurofilament light chain (NfL), and glial fibrillary acidic protein (GFAP) in cerebrospinal fluid (CSF) and plasma, have shown elevated levels in PSP compared to healthy controls but limited utility in distinguishing PSP from other neurodegenerative conditions^10–14^. Total tau (t-tau) levels are generally low in PSP^15–17^. Combining NfL, GFAP, and t-tau in a ratio may offer a better understanding of the interplay between axonal injury, astrocyte pathology, and tau function, while improving diagnostic value.

This study evaluates ^18^F-PI-2620 uptake in patients with PSP, investigating its associations with fluid biomarkers, brain volume, and cognitive performance across key brain regions. We hypothesized that subcortical tau accumulation would be associated with higher levels of NfL, GFAP, t-tau and their ratios in CSF and plasma, with regional brain volume loss and impaired cognition. Additionally, fluid biomarkers were compared against controls. Our findings aim to advance the role of tau-PET as a valuable biomarker in clinical trials, potentially improving diagnosis and treatment evaluation when combined with other biomarkers.

## Methods

### Patient enrolment and sample characteristics

Patients were recruited as part of the SEL003 clinical trial of sodium selenate for the treatment of PSP^18^. This cross-sectional, multicentre analysis included the baseline data collected from 28 patients with clinically diagnosed probable PSP (Richardson’s syndrome) from six clinical centres across Australia (Melbourne, Sydney, Brisbane, and Adelaide). Baseline data, including ^18^F-PI-2620 tau-PET, brain MRI, CSF, blood, and cognitive testing were obtained from July 2021 to August 2024.

Additionally, 10 age- and sex-matched controls, cognitively normal (CDRs=0, MMSE>25) and without known neurological disease, with available CSF and blood samples, were included from the Monash University Brain and Cognitive Health (BACH) cohort study for comparison. Diagnosis of PSP was made according to the Movement Disorders Society criteria for possible/probable PSP. Demographic data and full medical history were collected during screening. Inclusion criteria required participants to be over 40 years old, have symptom onset within five years, and live in the community with at least 10 contact hours per week with a caregiver and were free form other neurological disorders or structural brain MRI abnormalities. Informed written consent was obtained from all patients or their legal representatives, and from the caregiver. Ethics approval was granted by the Alfred Health and Monash University Human Research Ethics Committee, Melbourne (594/20, SEL003; HREC/69184/Alfred-2020; BACH, 532/21, AH). The SEL003 trial is registered with the Australian New Zealand Clinical Trials Registry (ACTRN12620001254987).

### CSF and blood analyses

Plasma (6 mL) and CSF (20 mL) samples from all participants were analysed using the single molecule array (SIMOA) platform (Quanterix Corp., Lexington, MA). The Neurology 4-Plex A kits were used to measure NfL, GFAP, and t-tau, following the manufacturer’s protocols. All samples were obtained fasting and frozen at -80°C and stored and analysed by the same lab. The coefficients of variance [range] were the following: CSF GFAP=5.02% [0.19-53.3%]; plasma GFAP=2.59% [0.07-11.3%]; CSF NfL=7.07% [0.23-10.7%]; plasma NfL=4.75% [1.1-8.6%]; CSF tau=2.72% [0.07-8.6%]; plasma tau=7.69% [0.03-30.1%]. Additional biomarker ratios (NfL/t-tau, GFAP/t-tau, GFAP/NfL) were calculated.

### MRI and PET acquisition and preprocessing

Whole-brain 3D T1-weighted 3T MRI images (0.8 mm isotropic voxels) and ^18^F-PI-2620 tau-PET (Life Molecular Imaging, Berlin, Germany) were acquired. PET scans followed a full dynamic protocol from 0 to 60 minutes post-injection (185 MBq ± 10%), with frames of 10×30s, 5×60s, and 10×300s. PET images were reconstructed using the VPFXS algorithm, with attenuation correction applied and a smoothing filter, resulting in voxel dimensions of 2.3×2.3×3.3 mm. PET and MRI image preprocessing steps are described in the **eMethods** in the **Supplement 1.**

### Cognitive testing

A comprehensive cognitive and symptom assessment was conducted, including the PSP Rating Scale (PSPRS), Behaviour Rating Inventory of Executive Function-Adult (BRIEF, ‘Never’ scale), Frontal Assessment Battery (FAB), Digit Span subtest from the Wechsler Adult Intelligence Scale – Fourth Edition (forwards and reverse trials), Trail Making Test (TMT A and B), the Category Fluency Test, Controlled Oral Word Association Test (COWAT), Victoria Stroop Test, and the Hayling Sentence Completion Test. A composite score was derived by averaging the min-max normalized scores of the Digit Span reverse trial, TMT-B (reversed scaling), and Stroop Interference score. Each test was scaled between 0 and 1 and higher composite scores indicated better performance.

### Data analysis

ROI-level statistical analyses were conducted in R (version 4.3.1), and voxel-wise analyses were performed using SPM12^19^ in Matlab (version R2022a). ROI-based analyses were tested at *p*<0.05 and voxel-wise analyses were tested at the *p*<0.001 significance level, 2-sided.

Differences in demographics and fluid biomarkers between PSP patients and controls were assessed using the Wilcoxon signed-rank and chi-squared tests, with effect sizes reported using rank-biserial correlation or Cramér’s V. ROC analyses were conducted to evaluate biomarker discrimination and optimal cut-off points (pROC package).

Linear mixed-effects models (lme4 package) were employed to investigate associations between tau uptake (dependent variable: SUVr), fluid biomarkers, brain volume, and cognitive test scores. Fixed effects included age, sex, disease duration and tau uptake region (pallidum, putamen, accumbens, red nucleus, subthalamic nucleus, substantia nigra, caudate, thalamus, frontal, temporal, parietal, posterior cingulate, anterior cingulate, and occipital). Random intercepts were specified for each participant to account for within-subject variability.

Separate models were fitted for each fluid biomarker type (NfL, GFAP, t-tau, their ratios) in both plasma and CSF to assess the association of biomarker level with tau uptake. An additional model evaluated the relationship between regional brain volume and tau uptake, including an interaction term between brain region and tau uptake region to explore how volume influences SUVr. To analyse the association between cognition and tau uptake, separate models were used for each neuropsychological test (PSPRS, BRIEF, FAB, Digit Span subtests, TMT A and B, Category Fluency, COWAT, Stroop, Hayling, composite score).

Partial correlation coefficients (ppcor package) were computed to examine the associations between regional/total brain volume and tau uptake, controlling for age, sex, and disease duration. Voxel-wise multiple regression analyses were performed to explore the relationship between tau-PET uptake, fluid biomarkers, and cognition. Additional analyses assessing the associations between fluid biomarkers and clinical scales are reported in the **eFigure 1** in **Supplement 1**.

## Results

The PSP cohort consisted of 28 participants (median [IQR] age, 66 [62-70] years; 12 females [43%]). Detailed demographics and test results are provided in the **eTable 1** in **Supplement 1**. A subset of 24 patients (median [IQR] age, 67 [62-71] years; 11 females [46%]) and 10 controls (median [IQR] age, 65 [62-68] years; 6 females [60%]) were included for fluid biomarker group analyses. Patients and controls did not differ based on age and sex (**table 1**).

**Table 1.** Fluid biomarkers (pg/mL) and demographics in controls and a subset of PSP

| | | Controls (n=10) | PSP (n=24) | $r_{rb}^a$ | P value |
| --- | --- | --- | --- | --- | --- |
| Age, years, Mdn (IQR) |  | 65 (5.4) | 67 (8.2) | 0.14 | .70 |
| Sex, n (%) | Female | 6 (60) | 11 (46) | 0.06<sup>a</sup> | .71 |
|  | Male | 4 (40) | 13 (54) |  |  |
| NfL Mdn (IQR) | CSF | 505 (531) | 1567 (1524) | 0.58 | .001*** |
|  | Plasma | 14.5 (5.6) | 20.1 (13.4) | 0.4 | .02* |
| GFAP Mdn (IQR) | CSF | 8092 (5856) | 16128 (6853) | 0.49 | .006** |
|  | Plasma | 120 (79.1) | 170 (123) | 0.22 | .22 |
| t-tau Mdn (IQR) | CSF | 140 (69.0) | 110 (62.0) | 0.2 | .30 |
|  | Plasma | 3.6 (1.1) | 3.1 (2.1) | 0.23 | .21 |
| NfL/t-tau Mdn (IQR) | CSF | 4.7 (2.8) | 15 (7.0) | 0.74 | <.000*** |
|  | Plasma | 3.9 (1.2) | 7.2 (8.9) | 0.46 | .007** |
| GFAP/t-tau Mdn (IQR) | CSF | 72.5 (34.4) | 138 (74.2) | 0.59 | .001*** |
|  | Plasma | 42.6 (24.8) | 64.9 (51.7) | 0.41 | .02* |
| GFAP/NfL Mdn (IQR) | CSF | 15.1 (7.4) | 9.8 (7.2) | 0.41 | .02* |
|  | Plasma | 8.5 (4.0) | 8 (5.2) | 0.09 | .63 |

PSP=progressive supranuclear palsy, CSF=cerebrospinal fluid, NfL=neurofilament light chain,
GFAP=glial fibrillary acidic protein, tTau=total tau, Mdn=median, IQR=interquartile range. Asterisks
indicate significant Wilcoxon signed-rank tests. \* $p<0.05$ ; \*\* $p<0.01$ , \*\*\* $p<0.001$ . <sup>a</sup>Effect size using
rank-biserial correlation. <sup>b</sup>Effect size using Cramér's V strength of association of the Chi-square test.

### Subcortical tau uptake is associated with increased NfL and decreased GFAP or t-tau

ROI-based analyses revealed significant interaction effects between fluid biomarkers and regions of tau binding. Associations were primarily observed in subcortical areas such as the pallidum, putamen, red nucleus, substantia nigra, subthalamic nucleus, and to a lesser extent the accumbens. Significant negative associations were found for plasma and CSF GFAP/NfL (putamen: β=-0.02; 95% CI, -0.01--0.01; *p*<.001), plasma t-tau (substantia nigra: β=-0.05; 95% CI, -0.08--0.02; *p*=.001), plasma GFAP (subthalamic nuclei: β=-0.001; 95% CI, -0.000-0.000; *p*=.003), and a trend observed for CSF GFAP (substantia nigra: β<0.001; 95% CI, 0.000-0.000; *p*=.09). Positive associations were found for CSF NfL/t-tau (putamen: β=0.01; 95% CI, -0.01-0.02; *p*<.001), and plasma and CSF NfL (putamen: β<0.001; 95% CI, 0.000-0.000; *p*lll=.008). No significant associations were found for plasma or CSF GFAP/t-tau, nor for CSF t-tau (**Figure 1 A**). Detailed results are shown in the **eTable 2** in **Supplement 1.** Voxel-wise analysis show the significant association between plasma GFAP/NfL and tau uptake in the pallidum and putamen (*p<*.001, *k*>100) (**Figure 1 B**).

**Figure 1.**
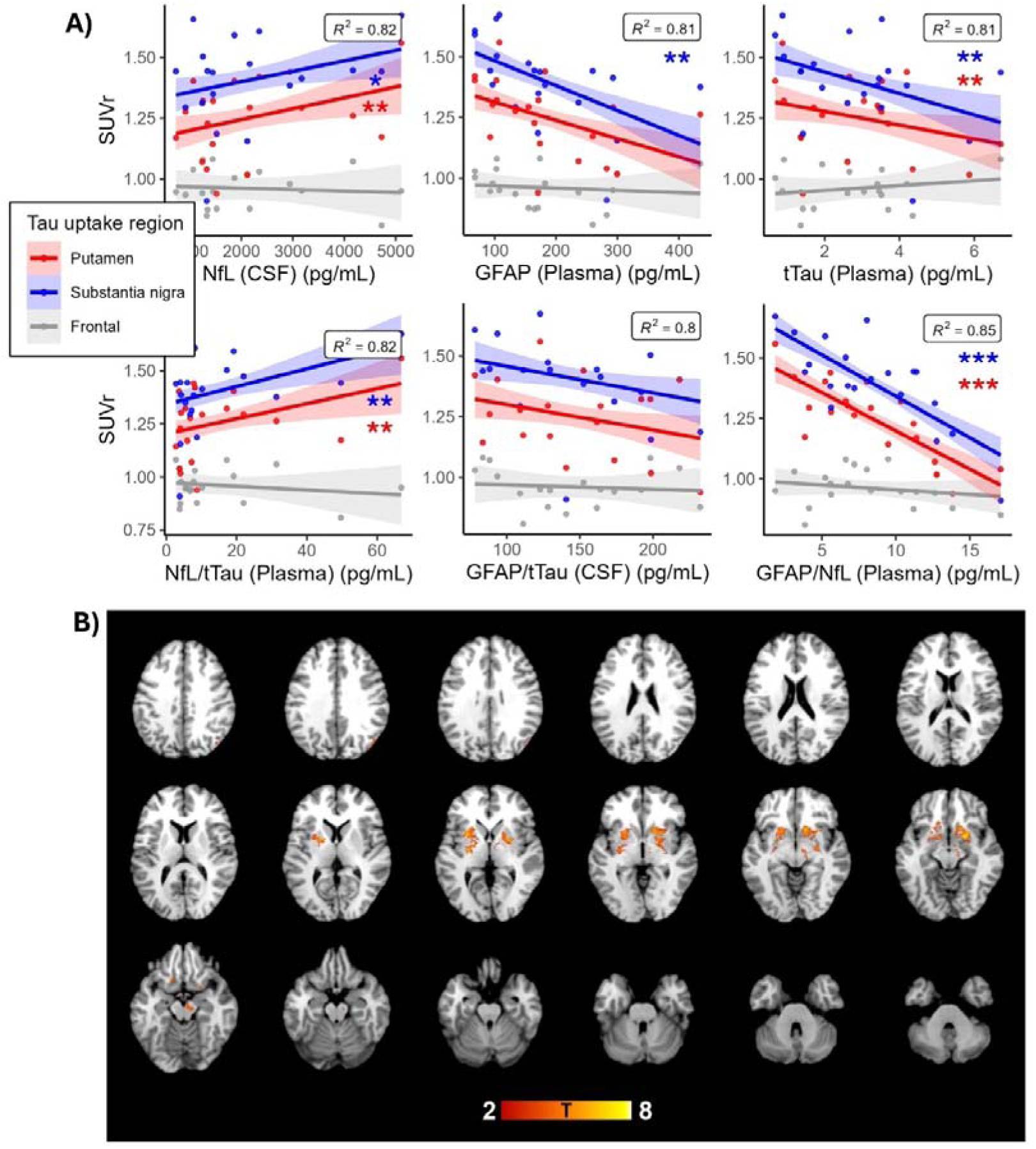
Relationship between tau uptake and fluid biomarker levels. A) ROI-based linear mixed effect models, with age, sex, and disease duration fixed effects, shown across 3 tau uptake brain regions (putamen, substantia nigra, frontal). \**p*<.05; \*\**p*<.01, \*\*\**p*<.001. Shaded areas represent 95% confidence intervals around fitted lines. B) Voxel-wise multiple regression relationship between tau uptake and plasma GFAP/NfL (pg/mL) in the basal ganglia, with T-statistics shown at *p<*.001, *k*>100, overlaid on a standard template T1 MRI image.

### Tau uptake is associated with total brain and basal ganglia atrophy

Significant negative correlations were found between total brain volume and tau uptake across the brain, including basal ganglia, midbrain, and cortex (*r*=0.68, *p*<.05) (**Figure 2 A**). Additional correlations were observed between putamen or pallidum volume and tau uptake across the same regions (*r*=0.7, *p*<.05), but fewer correlations with tau uptake in frontal regions or the red nucleus (*r*<0.4, *p*>.05).

**Figure 2.**
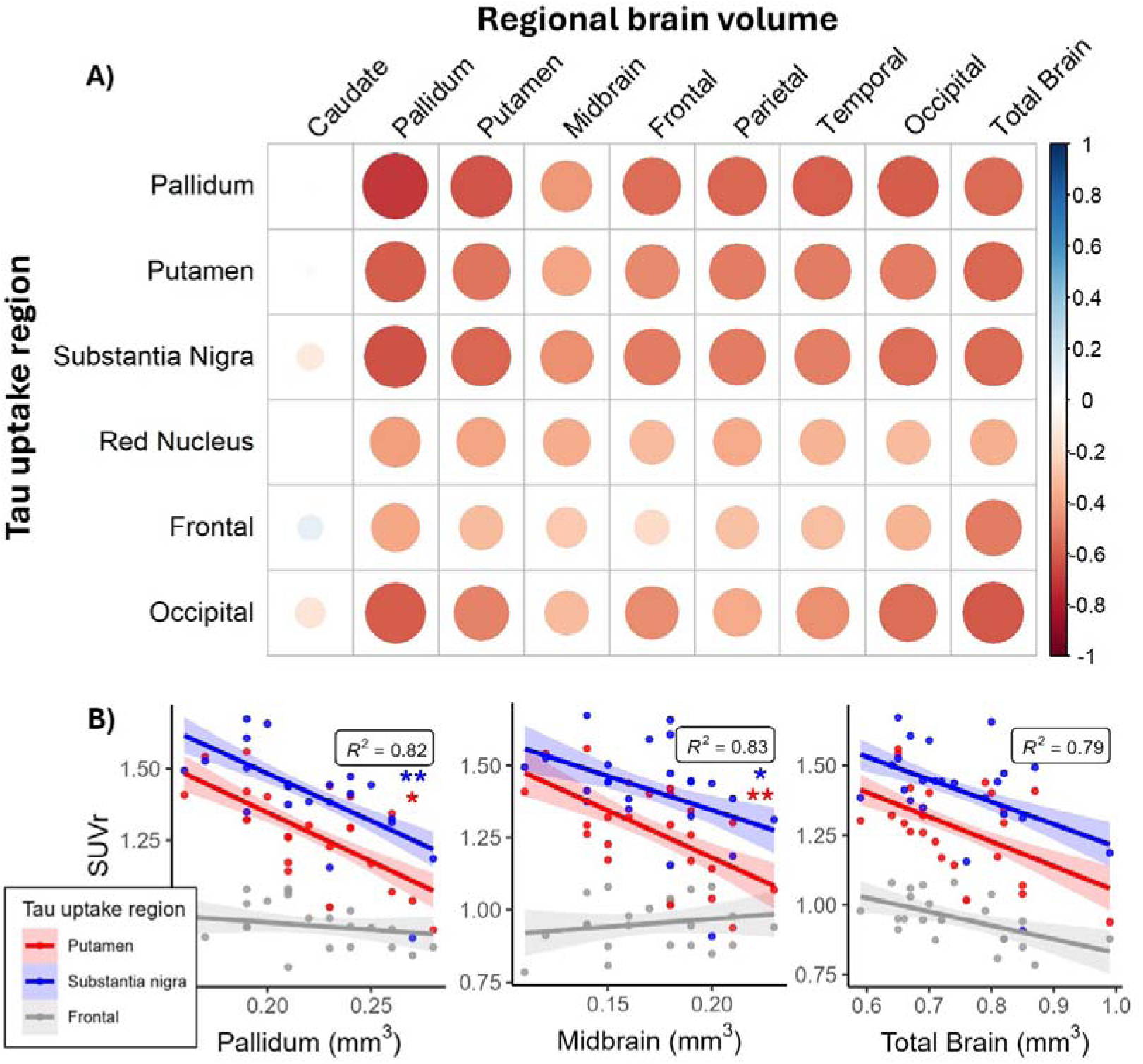
Relationship between tau uptake and volume. A) Partial correlation heatmap across brain regions of atrophy and tau uptake regions, based on spearman correlation analysis controlled for age, sex, and disease duration. B) Linear mixed effect models, with age, sex, and disease duration fixed effects, shown across 3 selected tau uptake regions (putamen, substantia nigra, frontal) shown for pallidum, midbrain, and total brain volume. \**p*<.05; \*\**p*<.01, \*\*\**p*<.001. Shaded areas represent 95% confidence intervals around fitted lines.

Further analysis revealed that lower volume explained variations in tau uptake (e.g. putamen: β=-0.97; 95% CI, -1.5--0.42; *p*=.001) (**Figure 2 B**). There were significant interactions between midbrain volume and subcortical tau uptake (e.g. putamen: β=-2.4; 95% CI, -4.1--0.73; *p*=.005), and between pallidum volume and subcortical tau uptake (e.g. putamen: β=-1.8; 95% CI, -3.5--0.23; *p*=.03).

### Tau uptake is associated with executive impairments and disease severity

Linear mixed-effects models revealed significant interaction effects between test scores and SUVr, indicating that the relationships between clinical performance and tau uptake varied across different brain regions. Significant negative associations were found for the composite score (putamen: β=-0.64; 95% CI, -1.04--0.24; *p*=.003), Digit Span reverse trial (putamen: β=-0.029; 95% CI, -0.05--0.01; *p*=.009), COWAT (red nucleus: β=-0.007; 95% CI, -0.01--0.00; *p*=.01), and BRIEF (red nucleus: β=-0.003; 95% CI, -0.01--0.00; *p*=.016). Positive associations were seen with TMT-A and B (putamen: β=0.002; 95% CI, 0.00-0.00; *p*=.008) (**Figure 3 A**). No significant associations were observed for the PSPRS, Digit Span forward, FAB, Stroop, Hayling, or Category Fluency Test. Detailed results are provided in **eTable 3** in **Supplement 1.** Voxel-wise analysis showed significant associations between tau uptake and executive function tasks (e.g., Digit Span reverse) in the pallidum and putamen (*p<*.01, *k*>50) (**Figure 3 B**).

**Figure 3.**
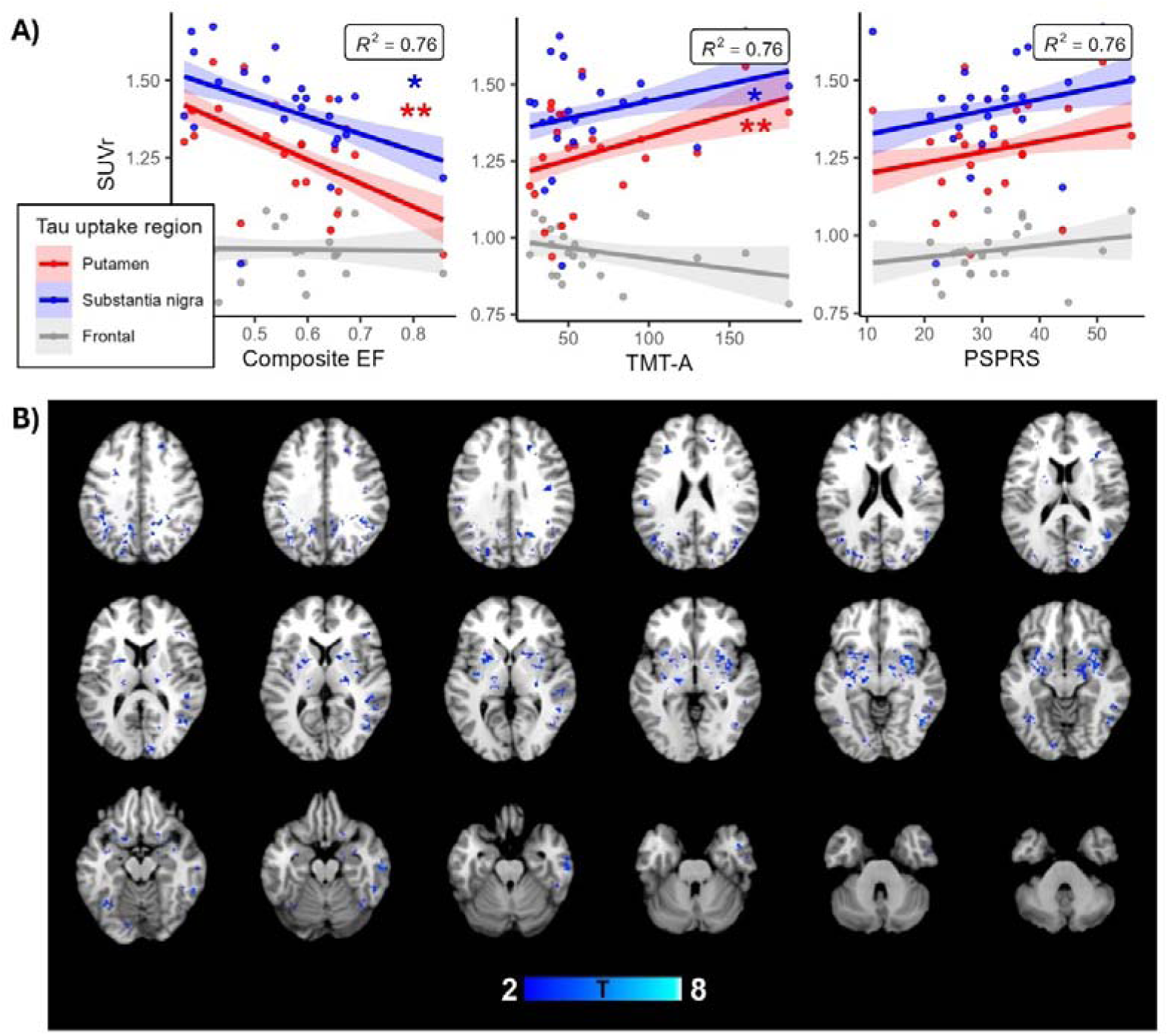
Relationship between tau uptake and clinical scales. A) ROI-based linear mixed effect models, with age, sex, and disease duration fixed effects, shown across 3 selected tau uptake brain regions (putamen, substantia nigra, frontal), shown for the composite executive function score (Stroop, Digit Span reverse, TMT-B), TMT-A, and PSPRS at \**p*<.05; \*\**p*<.01, \*\*\**p*<.001. Shaded areas represent 95% confidence intervals around fitted lines. B) Voxel-wise multiple regression relationship between tau uptake and performance on the Digit Span reverse, with T-statistics shown at *p<*.01, *k*>50, overlaid on a standard template T1 MRI image.

### Higher NfL and GFAP levels in PSP compared to controls

Patients with PSP displayed significantly higher levels of GFAP (CSF), NfL (CSF and plasma), but not t-tau, compared to controls. Biomarker ratios demonstrated larger group differences with higher levels of GFAP/t-tau (CSF and plasma), NfL/t-tau (CSF and plasma), and significantly lower levels of GFAP/NfL (CSF) in PSP as compared to controls (**Table 1** and **figure 4**).

**Figure 4.**
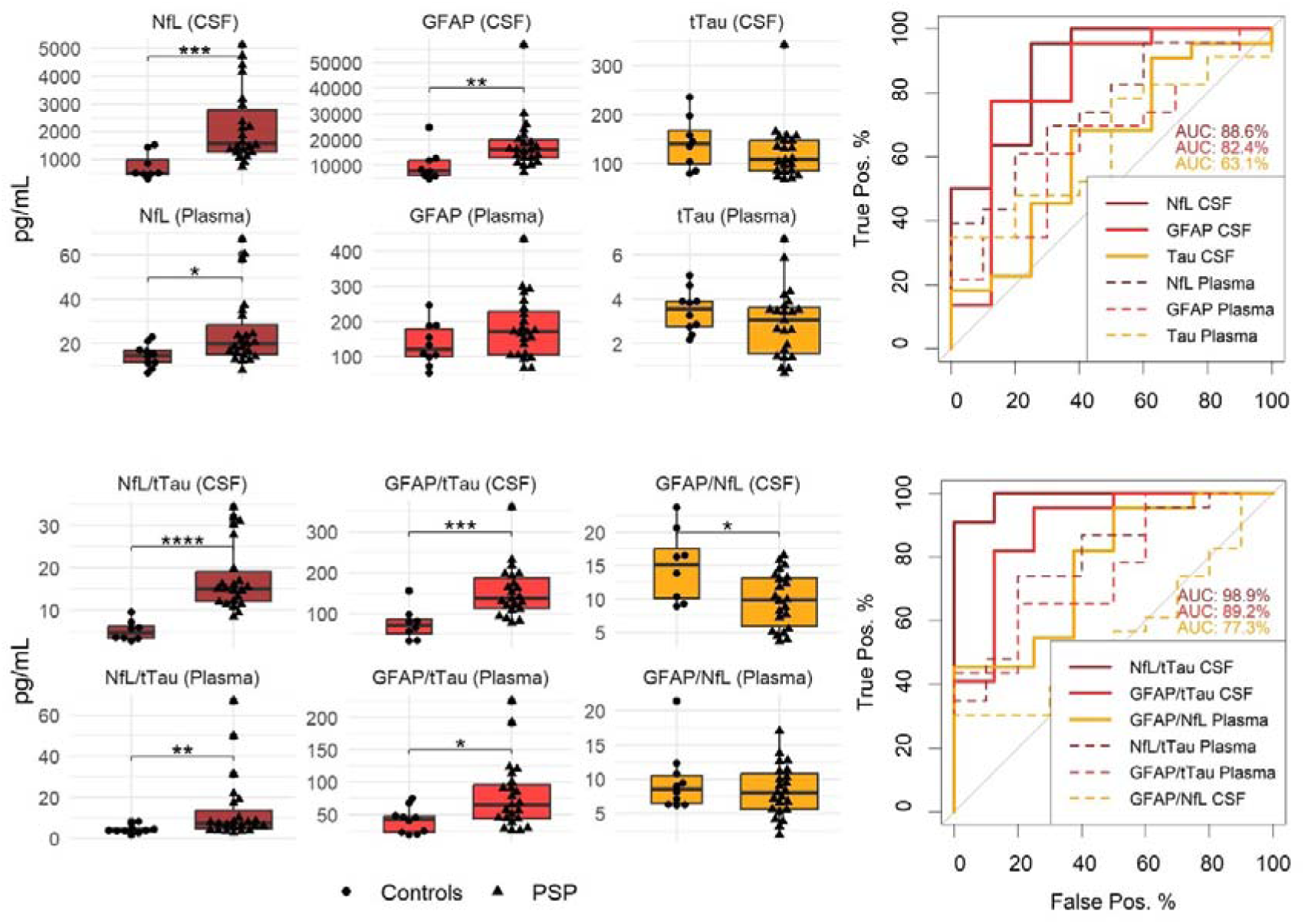
Fluid biomarker differences between groups and discriminatory power. . Vertical lines in boxplots represent error bars. Asterisks indicate significant Wilcoxon signed-rank tests. \**p*<.05; \*\**p*<.01, \*\*\**p*<.001. AUCs are shown for the stronger CSF results (thick lines). PSP=progressive supranuclear palsy, CSF=cerebrospinal fluid, NfL=neurofilament light chain, GFAP=glial fibrillary acidic protein, tTau=total tau.

Next, ROC analyses were performed. The AUC of CSF NfL/t-tau showed a high power of 99% (95% CI: 96%-100%), with an optimal cut-off of 11. CSF GFAP/t-tau differentiated the groups with an AUC of 89% (95% CI: 74%-100%) and a cut-off value of 84. The CSF GFAP/NfL ratio showed an AUC of 77% (95% CI: 58%-97%) with a cut-off of 3.9 pg/mL. The AUCs for plasma, as well as for NfL, GFAP, and t-tau, are reported in the **eResults** in the **Supplement 1** and shown in **figure 4**. Some clinical scales were associated with fluid biomarkers and are described in the **eFigure 1** in **Supplement 1**.

## Discussion

This cross-sectional study demonstrated that subcortical tau accumulation in PSP, as measured by ^18^F-PI-2620 tau-PET, is associated with fluid biomarkers of neuronal injury and inflammation, as well as with brain atrophy on MRI and poorer cognition. We observed significant differences in fluid biomarker levels, particularly in NfL, GFAP, and the ratios of CSF NfL/t-tau, GFAP/t-tau, when comparing patients with PSP to controls. These findings suggest that ^18^F-PI-2620 is a valuable tool for detecting pathology and its impact on neurodegeneration and clinical presentation, supporting its use as a biomarker endpoint for clinical trials and monitoring the effects of treatment. Additionally, NfL, GFAP, NfL/t-tau, GFAP/t-tau, and GFAP/NfL may serve as surrogate markers to aid in the diagnosis of PSP.

Consistent with previous research, ^18^F-PI-2620 showed high sensitivity for detecting tau accumulation in PSP^8,9,20^. Our study found associations in the globus pallidus and midbrain, aligning with post-mortem tau distribution patterns^2,4^ and prior in-vivo studies using ^18^F-PI-2620 in both PSP and CBS^8,9,20–22^. Neurodegenerative disease progression is thought to be driven by tau accumulation and its spread along functional brain networks^21,22^.

In PSP, elevated NfL levels in both CSF and blood, as observed in this study, are consistent with reports across several neurodegenerative diseases, including frontotemporal dementia, and likely indicate axonal damage and degeneration^23^. NfL levels are elevated in PSP and CBS relative to Parkinson’s disease and controls^12–14,24^, with baseline levels predictive of disease severity in PSP^12,25^. Although not specific to PSP, elevated NfL may reflect more aggressive neuronal loss in rapidly progressing conditions like PSP and frontotemporal dementia compared to Parkinson’s disease or Alzheimer’s disease^10,11,26^.

Interestingly, we found that plasma and CSF GFAP levels, a marker of astrocytic activation or loss, were elevated in PSP compared to controls, but lower GFAP levels were paradoxically associated with increased tau uptake and impaired cognition. It is possible that lower GFAP levels reflect impaired astrocytic function as the disease progresses, which needs further investigation, as GFAP is typically elevated in various neurological disorders, including PSP^10,11,14,27^.

Our findings also indicated that fluid biomarker ratios, such as NfL/t-tau and GFAP/t-tau, provide more disease-specific insights than individual biomarker levels. For example, the GFAP/NfL ratio may indicate disproportionate astrocytic activation relative to neuronal damage in PSP, serving as a potential marker for disease staging. This ratio has previously been identified as a marker specific to tauopathies^28^, and in our study, its strong association with tau-PET highlights its potential diagnostic utility. Although the GFAP/t-tau ratio was elevated in PSP compared to controls, it did not show associations with tau burden or cognition, suggesting that GFAP might serve as an early marker of disease before tau pathology manifests clinically.

Generally, our findings suggest that CSF measures, including NfL, GFAP, NfL/t-tau, GFAP/t-tau, and GFAP/NfL more robustly distinguish PSP from controls compared to blood measures. While blood-based biomarkers are less invasive and more accessible, they are prone to peripheral contamination and reduced signal strength compared to CSF markers.

In line with previous studies, t-tau levels in both CSF and plasma showed no significant differences between the groups, but a trend towards lower levels was observed in PSP, particularly in plasma. This aligns with reports of reduced t-tau in PSP compared to other tauopathies^15–17^. Moreover, CSF t-tau has been associated with decreased perfusion in subcortical areas in PSP^9^, suggesting that low t-tau could be a characteristic marker of PSP. Elevated t-tau levels are common in other neurological conditions^29–34^, making t-tau a general marker of neuronal damage. However, the observed lower t-tau levels in PSP might reflect processes specific to 4R-tau pathology.

We also found that tau-PET uptake in the basal ganglia, midbrain, and occipital regions was associated with widespread brain atrophy, particularly in the basal ganglia and total brain. The absence of significant associations between frontal tau uptake and atrophy might reflect the lack of tau accumulation in this region at this early disease stage. Additionally, the lack of association between caudate volume and tau uptake suggests that caudate atrophy may not be sensitive to tau load or atrophy. Previous studies indicated hypoperfusion in the caudate using ^18^F-PI-2620 in PSP as an indicator of neuronal damage^9,35^. However, it remains unclear whether perfusion deficits and structural damage are interchangeable. It is important to recognize that tau accumulation and neurodegeneration are distinct processes, and we propose that ^18^F-PI-2620 detects tau accumulation earlier than MRI can capture structural damage. However, this temporal relationship cannot be definitely established from this cross-sectional sectional analysis.

Our study also revealed that tau-PET uptake was associated with executive impairments and disease severity. A lack of correlation with the PSPRS scale in this study may be attributed to the early disease stage represented in this cohort. However, our findings suggest that tau-PET may be useful in evaluating cognitive impairment in PSP, as previous studies had not assessed ^18^F-PI-2620 in combination with comprehensive cognitive batteries, or did not find significant associations^8^.

A limitation of this study is the lack of autopsy confirmation for PSP diagnoses, which may have introduced diagnostic inaccuracies. However, the homogeneous sample of PSP patients with Richardson’s Syndrome minimizes potential confounding effects from different subtypes. While ^18^F-PI-2620 demonstrates promising results, we cannot entirely rule out off-target binding to Monoamine oxidase A and B (MAO-A and -B), a known limitation of first-generation tau tracers^36–39^. Among second-generation tau-PET tracers, only ^18^F-PM-PBB3 has shown some promise in detecting primary tauopathies, however, larger sample sizes and autopsy validation studies are still needed^40,41^. Additionally, biomarkers such as NfL, GFAP, and t-tau are not specific to tau pathology or PSP, serving as general markers of neuronal damage and inflammation. Fluid biomarkers do not provide region-specific information, which limits their utility in staging disease severity. We propose using these fluid biomarkers as potential surrogate markers alongside tau-PET. Newer markers such as MTBR-tau_243_ in CSF show promise as potential tau-specific biomarkers and should be explored further^42^. Future studies should also consider diurnal fluctuations in biomarker levels, as observed with Aβ in Alzheimer’s disease^43^. In our study plasma was obtained in the morning, while CSF was mostly obtained after lunch.

## Conclusion

This cross-sectional study highlights the utility of ^18^F-PI-2620 tau-PET in detecting subcortical tau pathology in PSP and its association with fluid biomarkers, brain volume, and cognition. The integration of tau-PET and fluid biomarkers could improve the diagnostic workflows and advance the development of targeted therapies for PSP and related tauopathies by using these biomarkers as reliable endpoints in clinical trials. Future research should focus on validating these biomarkers in longitudinal cohort studies and assessing their utility for early diagnosis.

## Data Availability

Data is part of an ongoing randomized controlled trial, thus will not be available immediately. Data will be available in the future following reasonable academic requests and ethics approval. Enquires should be directed to the corresponding author.

## Acknowledgements

**Author Contributions:** RD and LV had full access to all of the data in the study and take responsibility for the integrity of the data and the accuracy of the data analysis.

*Concept and design*, and interpretation of results: RD, LV, CM, KB, CD, TOB.

*Data acquisition*: RD, LV, CD, WOB.

Statistical analysis and drafting of the manuscript : RD.

Critical review of the manuscript for important intellectual content : LV, CM, WOB, SMD, KB, CD, MP, ML, TOB.

Administrative, technical, or material support: WOB, SMD, CD, KB, MP, ML, TOB, LV

*Supervision:*LV, CM, TOB.

All authors approve and agree to be accountable for all aspects of the work in ensure that questions related to the accuracy or integrity of any part of the work are appropriately investigated and resolved.

## Additional Contributions

We thank all the participants and controls for their time and participation in the study.

