## supplementary materials for "Linking PI-2620 tau-PET, fluid biomarkers, MRI, and cognition to advance diagnostics for progressive supranuclear palsy"

**Supplement 1**

**eMethods**

PET and MRI images were preprocessed using FSL (version 6.0.7.10). Dynamic PET images were motion-corrected using rigid alignment and averaged to create a single static PET image. The static PET image was co-registered to the corresponding T1-weighted MRI using linear transformation and then normalized to MNI152 through non-linear warping. Late-phase standardized uptake value ratio SUVr images (20-40 minute post-injection) were generated using cerebellum grey matter as the reference region to assess tau-specific PET uptake. SUVr maps were used for voxel-wise analyses and region of interest (ROI) analyses based on the Brainnetome^1^ atlas (pallidum, putamen, accumbens, caudate, thalamus, frontal, temporal, parietal, posterior cingulate, anterior cingulate, occipital region) and from the Talairach^2^ atlases for midbrain regions (red nucleus, subthalamic nucleus, substantia nigra). For volumetric analyses, MNI-normalized T1-weighted images were corrected for intracranial volume and segmented using the same atlases for the pallidum, putamen, caudate, whole midbrain, frontal, occipital, parietal, temporal, and whole brain volume.

**eResults**

ROC analyses were performed to explore the discriminatory power of biomarkers and their optimal cut-off points. The AUC for CSF NfL/t-tau, GFAP/t-tau, and GFAP/NfL are reported in the main text. The AUC of CSF Nfl was 89% (95% CI: 74%-100%), with an optimal cut-off of 911 pg/mL, of CSF GFAP was 82% (95% CI: 61%-100%), with an optimal cut-off of 13077 pg/mL, of CSF t-tau was 63% (95% CI: 39%-88%), of plasma NfL was 76% (95% CI: 58%-93%), of plasma GFAP was 64% (95% CI: 43%-84%), plasma t-tau was 64% (95% CI: 45%-84%), of plasma NfL/t-tau was 80% (95% CI: 63%-96%), of plasma GFAP/t-tau was 76% (95% CI: 58%-94%), and of plasma GFAP/NfL was 56% (95% CI: 35%-76%).

**eTables**

**eTable 1.** Demographics and average test results in the whole PSP cohort

|  | **PSP (n=28)** | |
| --- | --- | --- |
| Age, Mdn (IQR) | 66 | (8) |
| Female, n (%) | 12 | (43) |
| Male, n (%) | 16 | (57) |
| disease duration, Mdn weeks (IQR) | 50 | (204) |
| PSPRS, Mdn (IQR) | 31 | (10.2) |
| BRIEF, Mdn (IQR) | 34 | (27.5) |
| Category Fluency, Mdn (IQR) | 13 | (5.2) |
| Digit Span forward, Mdn (IQR) | 10 | (3.0) |
| Digit Span reverse, Mdn (IQR) | 7 | (2.0) |
| FAB, Mdn (IQR) | 14 | (3.2) |
| Hayling, Mdn (IQR) | 4 | (1.9) |
| Stroop, Mdn (IQR) | 12 | (4.5) |
| TMT-A, Mdn (IQR) | 50 | (44.6) |
| TMT-B, Mdn (IQR) | 180 | (100) |
| COWAT, Mdn (SD) | 17 | (8.2) |

PSPRS= PSP Rating Scale, FAB=Frontal Assessment Battery, TMT=Trail Making Test, COWAT=Controlled Oral Word Association Test, EF=executive function.

**eTable 2:** Relationships between tau uptake and fluid biomarkers (pg/mL) across brain regions

|  |  |  | **Estimate (β)** | **(95 % CI)** | | ***p*** |
| --- | --- | --- | --- | --- | --- | --- |
| **NfL** | CSF | Pallidum | 0.000 | (0.00 | 0.00) | 0.029* |
|  |  | Putamen | 0.000 | (0.00 | 0.00) | 0.008** |
|  |  | Accumbens | 0.000 | (0.00 | 0.00) | 0.056 |
|  |  | Caudate | 0.000 | (0.00 | 0.00) | 0.149 |
|  |  | Subst. Nigra | 0.000 | (0.00 | 0.00) | 0.01* |
|  |  | Subthal. Nucleus | 0.000 | (0.00 | 0.00) | 0.092 |
|  |  | Red Nucleus | 0.000 | (0.00 | 0.00) | 0.006** |
|  |  | Frontal | 0.000 | (0.00 | 0.00) | 0.769 |
|  |  | Occipital | 0.000 | (0.00 | 0.00) | 0.376 |
|  |  | Temporal | 0.000 | (0.00 | 0.00) | 0.476 |
|  |  | PCC | 0.000 | (0.00 | 0.00) | 0.828 |
|  |  | ACC | -0.000 | (0.00 | 0.00) | 0.851 |
|  |  | Thalamus | -0.000 | (0.00 | 0.00) | 0.939 |
|  | Plasma | Pallidum | 0.003 | (0.00 | 0.01) | 0.095 |
|  |  | Putamen | 0.004 | (0.00 | 0.01) | 0.023* |
|  |  | Accumbens | 0.002 | (0.00 | 0.01) | 0.119 |
|  |  | Caudate | 0.001 | (-0.00 | 0.00) | 0.452 |
|  |  | Subst. Nigra | 0.003 | (0.00 | 0.01) | 0.045* |
|  |  | Subthal. Nucleus | 0.002 | (-0.00 | 0.01) | 0.305 |
|  |  | Red Nucleus | 0.003 | (0.00 | 0.01) | 0.052 |
|  |  | Frontal | 0.000 | (-0.00 | 0.00) | 0.762 |
|  |  | Occipital | 0.001 | (-0.00 | 0.00) | 0.443 |
|  |  | Temporal | 0.001 | (-0.00 | 0.00) | 0.502 |
|  |  | PCC | 0.000 | (-0.00 | 0.00) | 0.798 |
|  |  | ACC | -0.001 | (-0.01 | 0.00) | 0.47 |
|  |  | Thalamus | -0.001 | (-0.00 | 0.00) | 0.617 |
| **GFAP** | CSF | Pallidum | -0.000 | (0.00 | 0.00) | 0.179 |
|  |  | Putamen | -0.000 | (0.00 | 0.00) | 0.102 |
|  |  | Accumbens | -0.000 | (0.00 | 0.00) | 0.487 |
|  |  | Caudate | 0.000 | (0.00 | 0.00) | 0.951 |
|  |  | Subst. Nigra | -0.000 | (0.00 | 0.00) | 0.086 |
|  |  | Subthal. Nucleus | -0.000 | (0.00 | 0.00) | 0.09 |
|  |  | Red Nucleus | 0.000 | (0.00 | 0.00) | 0.807 |
|  |  | Frontal | 0.000 | (0.00 | 0.00) | 0.684 |
|  |  | Occipital | -0.000 | (0.00 | 0.00) | 0.969 |
|  |  | Temporal | -0.000 | (0.00 | 0.00) | 0.952 |
|  |  | PCC | -0.000 | (0.00 | 0.00) | 0.882 |
|  |  | ACC | -0.000 | (0.00 | 0.00) | 0.661 |
|  |  | Thalamus | -0.000 | (0.00 | 0.00) | 0.662 |
|  | Plasma | Pallidum | -0.001 | (-0.00 | 0.00) | 0.069 |
|  |  | Putamen | -0.001 | (-0.00 | 0.00) | 0.069 |
|  |  | Accumbens | -0.000 | (-0.00 | 0.00) | 0.19 |
|  |  | Caudate | -0.000 | (-0.00 | 0.00) | 0.318 |
|  |  | Subst. Nigra | -0.001 | (-0.00 | 0.00) | 0.006** |
|  |  | Subthal. Nucleus | -0.001 | (-0.00 | 0.00) | 0.003** |
|  |  | Red Nucleus | -0.000 | (-0.00 | 0.00) | 0.138 |
|  |  | Frontal | 0.000 | (0.00 | 0.00) | 0.602 |
|  |  | Occipital | -0.000 | (-0.00 | 0.00) | 0.902 |
|  |  | Temporal | -0.000 | (-0.00 | 0.00) | 0.787 |
|  |  | PCC | -0.000 | (-0.00 | 0.00) | 0.761 |
|  |  | ACC | -0.000 | (-0.00 | 0.00) | 0.288 |
|  |  | Thalamus | -0.000 | (-0.00 | 0.00) | 0.225 |
| **tTau** | CSF | Pallidum | -0.000 | (-0.00 | 0.00) | 0.651 |
|  |  | Putamen | -0.000 | (-0.00 | 0.00) | 0.612 |
|  |  | Accumbens | -0.000 | (-0.00 | 0.00) | 0.675 |
|  |  | Caudate | 0.000 | (-0.00 | 0.00) | 0.617 |
|  |  | Subst. Nigra | -0.000 | (-0.00 | 0.00) | 0.777 |
|  |  | Subthal. Nucleus | -0.000 | (-0.00 | 0.00) | 0.793 |
|  |  | Red Nucleus | 0.000 | (-0.00 | 0.00) | 0.817 |
|  |  | Frontal | 0.000 | (-0.00 | 0.00) | 0.948 |
|  |  | Occipital | 0.000 | (-0.00 | 0.00) | 0.952 |
|  |  | Temporal | -0.000 | (-0.00 | 0.00) | 0.899 |
|  |  | PCC | -0.000 | (-0.00 | 0.00) | 0.851 |
|  |  | ACC | 0.000 | (-0.00 | 0.00) | 0.631 |
|  |  | Thalamus | -0.000 | (-0.00 | 0.00) | 0.743 |
|  | Plasma | Pallidum | -0.020 | (-0.05 | 0.01) | 0.223 |
|  |  | Putamen | -0.037 | (-0.07 | -0.01) | 0.024* |
|  |  | Accumbens | -0.024 | (-0.05 | 0.01) | 0.145 |
|  |  | Caudate | -0.006 | (-0.04 | 0.03) | 0.736 |
|  |  | Subst. Nigra | -0.053 | (-0.08 | -0.02) | 0.001** |
|  |  | Subthal. Nucleus | -0.043 | (-0.07 | -0.01) | 0.009** |
|  |  | Red Nucleus | -0.026 | (-0.06 | 0.00) | 0.109 |
|  |  | Frontal | 0.001 | (-0.03 | 0.03) | 0.951 |
|  |  | Occipital | -0.006 | (-0.04 | 0.02) | 0.693 |
|  |  | Temporal | -0.007 | (-0.04 | 0.02) | 0.668 |
|  |  | PCC | -0.018 | (-0.05 | 0.01) | 0.277 |
|  |  | ACC | 0.013 | (-0.02 | 0.05) | 0.524 |
|  |  | Thalamus | -0.004 | (-0.03 | 0.03) | 0.829 |
| **NfL/tTau** | CSF | Pallidum | 0.011 | (0.01 | 0.02) | 0.001** |
|  |  | Putamen | 0.013 | (0.01 | 0.02) | 0*** |
|  |  | Accumbens | 0.009 | (0.00 | 0.02) | 0.011* |
|  |  | Caudate | 0.005 | (-0.00 | 0.01) | 0.134 |
|  |  | Subst. Nigra | 0.010 | (0.00 | 0.02) | 0.003** |
|  |  | Subthal. Nucleus | 0.007 | (0.00 | 0.01) | 0.039* |
|  |  | Red Nucleus | 0.010 | (0.00 | 0.02) | 0.005** |
|  |  | Frontal | 0.001 | (-0.01 | 0.01) | 0.699 |
|  |  | Occipital | 0.003 | (-0.00 | 0.01) | 0.343 |
|  |  | Temporal | 0.003 | (-0.00 | 0.01) | 0.409 |
|  |  | PCC | 0.001 | (-0.01 | 0.01) | 0.684 |
|  |  | ACC | -0.001 | (-0.01 | 0.01) | 0.725 |
|  |  | Thalamus | -0.001 | (-0.01 | 0.01) | 0.855 |
|  | Plasma | Pallidum | 0.003 | (0.00 | 0.01) | 0.035* |
|  |  | Putamen | 0.004 | (0.00 | 0.01) | 0.005** |
|  |  | Accumbens | 0.003 | (0.00 | 0.01) | 0.051 |
|  |  | Caudate | 0.001 | (-0.00 | 0.00) | 0.342 |
|  |  | Subst. Nigra | 0.005 | (0.00 | 0.01) | 0.001** |
|  |  | Subthal. Nucleus | 0.003 | (0.00 | 0.01) | 0.026* |
|  |  | Red Nucleus | 0.005 | (0.00 | 0.01) | 0.002** |
|  |  | Frontal | 0.000 | (-0.00 | 0.00) | 0.998 |
|  |  | Occipital | 0.001 | (-0.00 | 0.00) | 0.522 |
|  |  | Temporal | 0.001 | (-0.00 | 0.00) | 0.553 |
|  |  | PCC | 0.001 | (-0.00 | 0.00) | 0.725 |
|  |  | ACC | -0.001 | (-0.00 | 0.00) | 0.708 |
|  |  | Thalamus | 0.000 | (-0.00 | 0.00) | 0.922 |
| **GFAP/tTau** | CSF | Pallidum | -0.000 | (-0.00 | 0.00) | 0.636 |
|  |  | Putamen | -0.001 | (-0.00 | 0.00) | 0.384 |
|  |  | Accumbens | 0.000 | (-0.00 | 0.00) | 0.938 |
|  |  | Caudate | -0.000 | (-0.00 | 0.00) | 0.628 |
|  |  | Subst. Nigra | -0.001 | (-0.00 | 0.00) | 0.34 |
|  |  | Subthal. Nucleus | -0.001 | (-0.00 | 0.00) | 0.248 |
|  |  | Red Nucleus | -0.000 | (-0.00 | 0.00) | 0.775 |
|  |  | Frontal | 0.000 | (-0.00 | 0.00) | 0.583 |
|  |  | Occipital | 0.000 | (-0.00 | 0.00) | 0.949 |
|  |  | Temporal | 0.000 | (-0.00 | 0.00) | 0.78 |
|  |  | PCC | 0.000 | (-0.00 | 0.00) | 0.692 |
|  |  | ACC | -0.001 | (-0.00 | 0.00) | 0.379 |
|  |  | Thalamus | -0.000 | (-0.00 | 0.00) | 0.726 |
|  | Plasma | Pallidum | -0.000 | (-0.00 | 0.00) | 0.933 |
|  |  | Putamen | 0.000 | (-0.00 | 0.00) | 0.624 |
|  |  | Accumbens | 0.000 | (-0.00 | 0.00) | 0.655 |
|  |  | Caudate | -0.000 | (-0.00 | 0.00) | 0.608 |
|  |  | Subst. Nigra | 0.001 | (0.00 | 0.00) | 0.264 |
|  |  | Subthal. Nucleus | 0.000 | (-0.00 | 0.00) | 0.791 |
|  |  | Red Nucleus | 0.000 | (-0.00 | 0.00) | 0.444 |
|  |  | Frontal | 0.000 | (-0.00 | 0.00) | 0.626 |
|  |  | Occipital | 0.000 | (-0.00 | 0.00) | 0.746 |
|  |  | Temporal | 0.000 | (-0.00 | 0.00) | 0.729 |
|  |  | PCC | 0.000 | (-0.00 | 0.00) | 0.545 |
|  |  | ACC | -0.001 | (-0.00 | 0.00) | 0.217 |
|  |  | Thalamus | -0.000 | (-0.00 | 0.00) | 0.462 |
| **GFAP/NfL** | CSF | Pallidum | -0.019 | (-0.03 | -0.01) | 0.002** |
|  |  | Putamen | -0.023 | (-0.03 | -0.01) | 0*** |
|  |  | Accumbens | -0.015 | (-0.03 | -0.00) | 0.014* |
|  |  | Caudate | -0.009 | (-0.02 | 0.00) | 0.134 |
|  |  | Subst. Nigra | -0.022 | (-0.03 | -0.01) | 0*** |
|  |  | Subthal. Nucleus | -0.018 | (-0.03 | -0.01) | 0.003** |
|  |  | Red Nucleus | -0.014 | (-0.03 | -0.00) | 0.02* |
|  |  | Frontal | -0.001 | (-0.01 | 0.01) | 0.86 |
|  |  | Occipital | -0.006 | (-0.02 | 0.01) | 0.297 |
|  |  | Temporal | -0.005 | (-0.02 | 0.01) | 0.406 |
|  |  | PCC | -0.001 | (-0.01 | 0.01) | 0.857 |
|  |  | ACC | -0.004 | (-0.02 | 0.01) | 0.527 |
|  |  | Thalamus | 0.002 | (-0.01 | 0.01) | 0.719 |
|  | Plasma | Pallidum | -0.023 | (-0.03 | -0.01) | 0*** |
|  |  | Putamen | -0.028 | (-0.04 | -0.02) | 0*** |
|  |  | Accumbens | -0.019 | (-0.03 | -0.01) | 0.001** |
|  |  | Caudate | -0.011 | (-0.02 | 0.00) | 0.075 |
|  |  | Subst. Nigra | -0.030 | (-0.04 | -0.02) | 0*** |
|  |  | Subthal. Nucleus | -0.027 | (-0.04 | -0.02) | 0*** |
|  |  | Red Nucleus | -0.020 | (-0.03 | -0.01) | 0.001** |
|  |  | Frontal | 0.000 | (-0.01 | 0.01) | 0.986 |
|  |  | Occipital | -0.005 | (-0.02 | 0.01) | 0.385 |
|  |  | Temporal | -0.006 | (-0.02 | 0.01) | 0.353 |
|  |  | PCC | -0.004 | (-0.02 | 0.01) | 0.521 |
|  |  | ACC | -0.005 | (-0.02 | 0.01) | 0.376 |
|  |  | Thalamus | 0.001 | (-0.01 | 0.01) | 0.908 |

NfL=neurofilament light chain, GFAP=glial fibrillary acidic protein, tTau=total tau. Asterisks indicate significant effects on linear mixed effect models, with age, sex, and disease duration fixed effects, **p*<0.05; ***p*<0.01, ****p*<0.001.

**eTable 3** Relationships between tau uptake and cognitive/behavioural scales across brain regions

|  |  | **Estimate (β)** | **(95 % CI)** | | ***p*** |
| --- | --- | --- | --- | --- | --- |
| **Digit Span Forward** | Pallidum | -0.014 | (-0.04 | 0.01) | 0.25 |
|  | Putamen | -0.015 | (-0.04 | 0.01) | 0.21 |
|  | Accumbens | -0.003 | (-0.03 | 0.02) | 0.79 |
|  | Caudate | -0.017 | (-0.04 | 0.01) | 0.17 |
|  | Subst. Nigra | 0.008 | (-0.02 | 0.03) | 0.51 |
|  | Subthal. Nucleus | -0.001 | (-0.02 | 0.02) | 0.95 |
|  | Red Nucleus | -0.023 | (-0.05 | -0.00) | 0.05 |
|  | Frontal | 0.006 | (-0.02 | 0.03) | 0.63 |
|  | Occipital | 0.012 | (-0.01 | 0.04) | 0.31 |
|  | Temporal | 0.009 | (-0.01 | 0.03) | 0.48 |
|  | PCC | 0.006 | (-0.02 | 0.03) | 0.64 |
|  | ACC | -0.011 | (-0.03 | 0.01) | 0.31 |
|  | Thalamus | -0.013 | (-0.04 | 0.01) | 0.27 |
| **Digit Span Reverse** | Pallidum | -0.025 | (-0.05 | -0.01) | 0.021* |
|  | Putamen | -0.029 | (-0.05 | -0.01) | 0.009** |
|  | Accumbens | -0.016 | (-0.04 | 0.01) | 0.15 |
|  | Caudate | -0.013 | (-0.03 | 0.01) | 0.25 |
|  | Subst. Nigra | -0.016 | (-0.04 | 0.00) | 0.14 |
|  | Subthal. Nucleus | -0.020 | (-0.04 | 0.00) | 0.07 |
|  | Red Nucleus | -0.024 | (-0.05 | -0.00) | 0.026* |
|  | Frontal | 0.005 | (-0.02 | 0.03) | 0.62 |
|  | Occipital | 0.003 | (-0.02 | 0.02) | 0.81 |
|  | Temporal | 0.002 | (-0.02 | 0.02) | 0.88 |
|  | PCC | 0.002 | (-0.02 | 0.02) | 0.88 |
|  | ACC | -0.014 | (-0.03 | 0.00) | 0.15 |
|  | Thalamus | -0.005 | (-0.03 | 0.02) | 0.67 |
| **Stroop** | Pallidum | -0.006 | (-0.02 | 0.01) | 0.48 |
|  | Putamen | -0.009 | (-0.03 | 0.01) | 0.28 |
|  | Accumbens | -0.000 | (-0.02 | 0.02) | 0.98 |
|  | Caudate | -0.007 | (-0.02 | 0.01) | 0.43 |
|  | Subst. Nigra | -0.004 | (-0.02 | 0.01) | 0.62 |
|  | Subthal. Nucleus | -0.006 | (-0.02 | 0.01) | 0.49 |
|  | Red Nucleus | 0.000 | (-0.02 | 0.02) | 0.97 |
|  | Frontal | 0.003 | (-0.01 | 0.02) | 0.71 |
|  | Occipital | 0.005 | (-0.01 | 0.02) | 0.57 |
|  | Temporal | 0.007 | (-0.01 | 0.02) | 0.43 |
|  | PCC | 0.002 | (-0.01 | 0.02) | 0.81 |
|  | ACC | -0.009 | (-0.02 | 0.01) | 0.24 |
|  | Thalamus | -0.007 | (-0.02 | 0.01) | 0.42 |
| **TMT-A** | Pallidum | 0.001 | (0.00 | 0.00) | 0.023* |
|  | Putamen | 0.002 | (0.00 | 0.00) | 0.008** |
|  | Accumbens | 0.001 | (0.00 | 0.00) | 0.13 |
|  | Caudate | 0.001 | (-0.00 | 0.00) | 0.40 |
|  | Subst. Nigra | 0.001 | (0.00 | 0.00) | 0.036* |
|  | Subthal. Nucleus | 0.001 | (0.00 | 0.00) | 0.04* |
|  | Red Nucleus | 0.001 | (0.00 | 0.00) | 0.09 |
|  | Frontal | -0.000 | (-0.00 | 0.00) | 0.45 |
|  | Occipital | 0.001 | (0.00 | 0.00) | 0.25 |
|  | Temporal | 0.000 | (-0.00 | 0.00) | 0.51 |
|  | PCC | 0.000 | (-0.00 | 0.00) | 0.75 |
|  | ACC | -0.000 | (-0.00 | 0.00) | 0.66 |
|  | Thalamus | 0.000 | (-0.00 | 0.00) | 0.53 |
| **TMT-B** | Pallidum | 0.001 | (0.00 | 0.00) | 0.029* |
|  | Putamen | 0.001 | (0.00 | 0.00) | 0.012* |
|  | Accumbens | 0.001 | (0.00 | 0.00) | 0.11 |
|  | Caudate | 0.000 | (0.00 | 0.00) | 0.43 |
|  | Subst. Nigra | 0.001 | (0.00 | 0.00) | 0.032* |
|  | Subthal. Nucleus | 0.001 | (0.00 | 0.00) | 0.041* |
|  | Red Nucleus | 0.000 | (0.00 | 0.00) | 0.35 |
|  | Frontal | 0.000 | (-0.00 | 0.00) | 0.89 |
|  | Occipital | 0.000 | (0.00 | 0.00) | 0.24 |
|  | Temporal | 0.000 | (0.00 | 0.00) | 0.38 |
|  | PCC | 0.000 | (0.00 | 0.00) | 0.53 |
|  | ACC | -0.000 | (-0.00 | 0.00) | 0.25 |
|  | Thalamus | -0.000 | (-0.00 | 0.00) | 0.94 |
| **Hayling** | Pallidum | -0.012 | (-0.04 | 0.01) | 0.37 |
|  | Putamen | -0.021 | (-0.05 | 0.01) | 0.13 |
|  | Accumbens | -0.011 | (-0.04 | 0.01) | 0.42 |
|  | Caudate | -0.006 | (-0.03 | 0.02) | 0.67 |
|  | Subst. Nigra | -0.010 | (-0.04 | 0.02) | 0.44 |
|  | Subthal. Nucleus | -0.012 | (-0.04 | 0.01) | 0.37 |
|  | Red Nucleus | 0.008 | (-0.02 | 0.03) | 0.55 |
|  | Frontal | -0.006 | (-0.03 | 0.02) | 0.63 |
|  | Occipital | -0.007 | (-0.03 | 0.02) | 0.61 |
|  | Temporal | -0.003 | (-0.03 | 0.02) | 0.81 |
|  | PCC | -0.001 | (-0.03 | 0.02) | 0.96 |
|  | ACC | 0.011 | (-0.01 | 0.04) | 0.40 |
|  | Thalamus | 0.005 | (-0.02 | 0.03) | 0.70 |
| **FAS** | Pallidum | -0.000 | (-0.01 | 0.01) | 0.97 |
|  | Putamen | -0.000 | (-0.01 | 0.00) | 0.87 |
|  | Accumbens | 0.001 | (-0.00 | 0.01) | 0.66 |
|  | Caudate | 0.001 | (-0.00 | 0.01) | 0.73 |
|  | Subst. Nigra | -0.003 | (-0.01 | 0.00) | 0.25 |
|  | Subthal. Nucleus | -0.004 | (-0.01 | 0.00) | 0.14 |
|  | Red Nucleus | -0.007 | (-0.01 | -0.00) | 0.01* |
|  | Frontal | 0.001 | (-0.00 | 0.01) | 0.58 |
|  | Occipital | 0.003 | (-0.00 | 0.01) | 0.28 |
|  | Temporal | 0.002 | (-0.00 | 0.01) | 0.43 |
|  | PCC | -0.000 | (-0.01 | 0.01) | 0.92 |
|  | ACC | 0.000 | (-0.01 | 0.01) | 0.95 |
|  | Thalamus | -0.001 | (-0.01 | 0.00) | 0.56 |
| **Category Fluency Test** | Pallidum | -0.002 | (-0.01 | 0.00) | 0.35 |
|  | Putamen | -0.001 | (-0.01 | 0.00) | 0.56 |
|  | Accumbens | -0.001 | (-0.01 | 0.00) | 0.67 |
|  | Caudate | -0.003 | (-0.01 | 0.00) | 0.25 |
|  | Subst. Nigra | 0.001 | (-0.00 | 0.01) | 0.83 |
|  | Subthal. Nucleus | -0.000 | (-0.01 | 0.01) | 1.00 |
|  | Red Nucleus | -0.003 | (-0.01 | 0.00) | 0.30 |
|  | Frontal | -0.001 | (-0.01 | 0.00) | 0.63 |
|  | Occipital | -0.000 | (-0.01 | 0.00) | 0.86 |
|  | Temporal | -0.000 | (-0.01 | 0.00) | 0.85 |
|  | PCC | 0.001 | (-0.00 | 0.01) | 0.66 |
|  | ACC | 0.001 | (-0.00 | 0.01) | 0.83 |
|  | Thalamus | -0.000 | (-0.01 | 0.00) | 0.93 |
| **Composite EF** | Pallidum | -0.520 | (-0.92 | -0.12) | 0.014* |
|  | Putamen | -0.641 | (-1.04 | -0.24) | 0.003** |
|  | Accumbens | -0.269 | (-0.67 | 0.13) | 0.20 |
|  | Caudate | -0.302 | (-0.70 | 0.10) | 0.16 |
|  | Subst. Nigra | -0.437 | (-0.83 | -0.04) | 0.04* |
|  | Subthal. Nucleus | -0.480 | (-0.88 | -0.08) | 0.024* |
|  | Red Nucleus | -0.281 | (-0.68 | 0.12) | 0.19 |
|  | Frontal | 0.097 | (-0.30 | 0.49) | 0.65 |
|  | Occipital | -0.003 | (-0.40 | 0.39) | 0.99 |
|  | Temporal | 0.059 | (-0.34 | 0.46) | 0.78 |
|  | PCC | -0.004 | (-0.40 | 0.39) | 0.99 |
|  | ACC | -0.198 | (-0.55 | 0.16) | 0.31 |
|  | Thalamus | -0.166 | (-0.56 | 0.23) | 0.43 |
| **FAB** | Pallidum | -0.001 | (-0.02 | 0.02) | 0.92 |
|  | Putamen | -0.004 | (-0.02 | 0.01) | 0.62 |
|  | Accumbens | 0.002 | (-0.01 | 0.02) | 0.86 |
|  | Caudate | -0.002 | (-0.02 | 0.01) | 0.83 |
|  | Subst. Nigra | -0.002 | (-0.02 | 0.01) | 0.85 |
|  | Subthal. Nucleus | -0.002 | (-0.02 | 0.01) | 0.80 |
|  | Red Nucleus | -0.014 | (-0.03 | 0.00) | 0.10 |
|  | Frontal | 0.004 | (-0.01 | 0.02) | 0.63 |
|  | Occipital | 0.005 | (-0.01 | 0.02) | 0.58 |
|  | Temporal | 0.003 | (-0.01 | 0.02) | 0.76 |
|  | PCC | 0.002 | (-0.01 | 0.02) | 0.82 |
|  | ACC | 0.004 | (-0.01 | 0.02) | 0.64 |
|  | Thalamus | -0.006 | (-0.02 | 0.01) | 0.43 |
| **CGI** | Pallidum | -0.008 | (-0.04 | 0.03) | 0.66 |
|  | Putamen | -0.011 | (-0.05 | 0.02) | 0.55 |
|  | Accumbens | -0.015 | (-0.05 | 0.02) | 0.43 |
|  | Caudate | -0.012 | (-0.05 | 0.02) | 0.50 |
|  | Subst. Nigra | -0.019 | (-0.05 | 0.02) | 0.32 |
|  | Subthal. Nucleus | -0.008 | (-0.04 | 0.03) | 0.65 |
|  | Red Nucleus | -0.036 | (-0.07 | -0.00) | 0.06 |
|  | Frontal | -0.007 | (-0.04 | 0.03) | 0.70 |
|  | Occipital | 0.003 | (-0.03 | 0.04) | 0.89 |
|  | Temporal | -0.008 | (-0.04 | 0.03) | 0.66 |
|  | PCC | 0.001 | (-0.03 | 0.04) | 0.98 |
|  | ACC | 0.015 | (-0.02 | 0.05) | 0.39 |
|  | Thalamus | -0.019 | (-0.05 | 0.02) | 0.30 |
| **BRIEF-A - 'Never'** | Pallidum | -0.002 | (-0.00 | 0.00) | 0.13 |
|  | Putamen | -0.002 | (-0.01 | 0.00) | 0.06 |
|  | Accumbens | -0.001 | (-0.00 | 0.00) | 0.29 |
|  | Caudate | -0.002 | (-0.00 | 0.00) | 0.12 |
|  | Subst. Nigra | -0.001 | (-0.00 | 0.00) | 0.60 |
|  | Subthal. Nucleus | -0.001 | (-0.00 | 0.00) | 0.29 |
|  | Red Nucleus | -0.003 | (-0.01 | -0.00) | 0.016* |
|  | Frontal | 0.000 | (-0.00 | 0.00) | 0.80 |
|  | Occipital | 0.001 | (-0.00 | 0.00) | 0.59 |
|  | Temporal | -0.000 | (-0.00 | 0.00) | 0.90 |
|  | PCC | 0.000 | (-0.00 | 0.00) | 0.99 |
|  | ACC | -0.000 | (-0.00 | 0.00) | 0.94 |
|  | Thalamus | -0.001 | (-0.00 | 0.00) | 0.43 |
| **PSPRS** | Pallidum | 0.001 | (-0.00 | 0.01) | 0.62 |
|  | Putamen | 0.001 | (-0.00 | 0.01) | 0.73 |
|  | Accumbens | 0.001 | (-0.00 | 0.01) | 0.75 |
|  | Caudate | 0.000 | (-0.01 | 0.01) | 0.92 |
|  | Subst. Nigra | 0.001 | (-0.00 | 0.01) | 0.62 |
|  | Subthal. Nucleus | 0.003 | (-0.00 | 0.01) | 0.28 |
|  | Red Nucleus | 0.004 | (-0.00 | 0.01) | 0.11 |
|  | Frontal | -0.001 | (-0.01 | 0.00) | 0.82 |
|  | Occipital | -0.000 | (-0.01 | 0.01) | 0.94 |
|  | Temporal | -0.000 | (-0.01 | 0.01) | 0.98 |
|  | PCC | -0.000 | (-0.01 | 0.01) | 0.99 |
|  | ACC | 0.002 | (-0.00 | 0.01) | 0.53 |
|  | Thalamus | -0.002 | (-0.01 | 0.00) | 0.35 |

**eFigures**

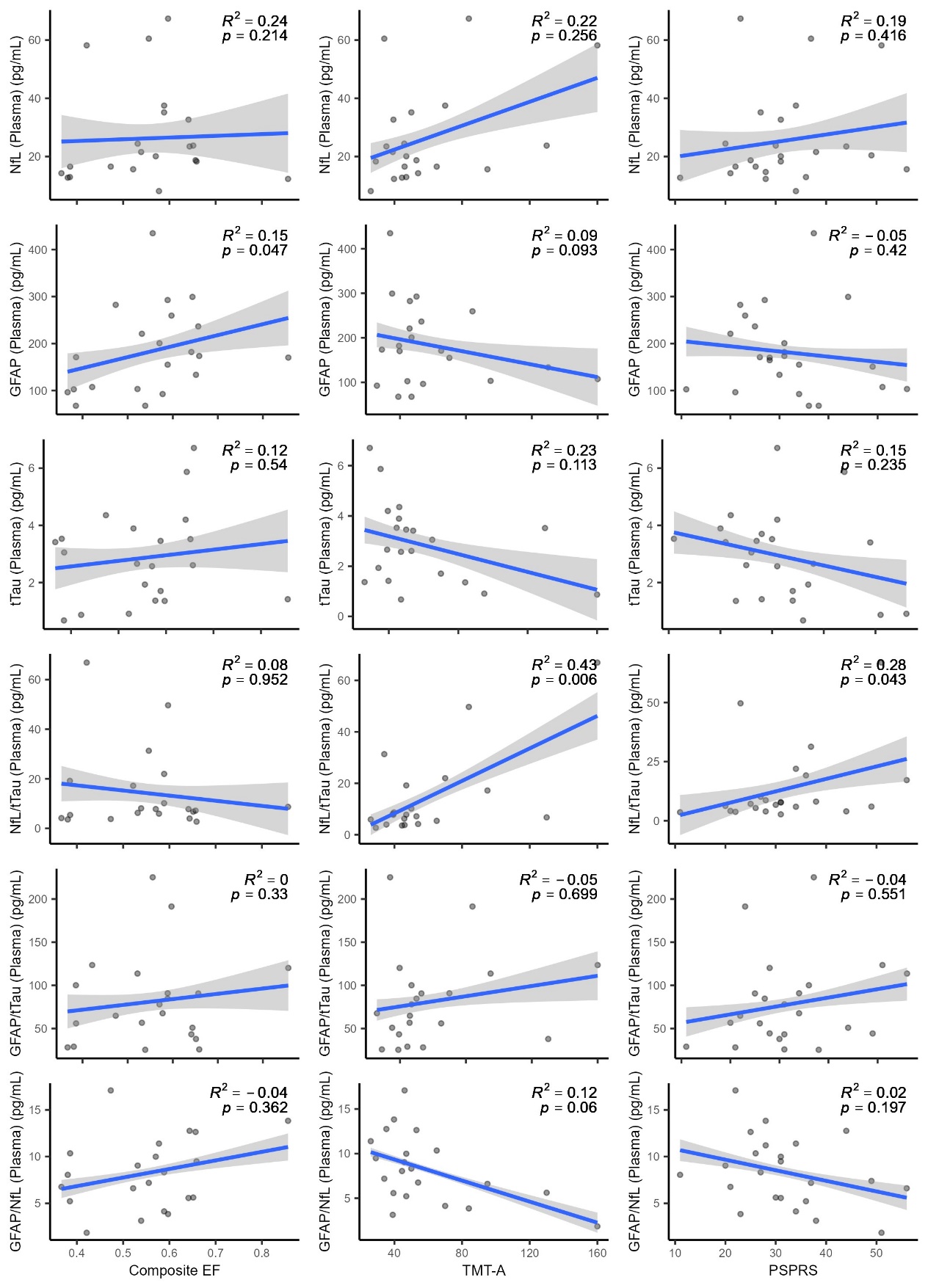

**eFigure 1. Relationship between plasma biomarkers and clinical scales.** ROI-based linear mixed effect models, with age, sex, and disease duration covariates.
